# Machine Learning techniques for the diagnosis of Schizophrenia based on Event Related Potentials

**DOI:** 10.1101/2022.03.24.22272090

**Authors:** Elsa Santos Febles, Marlis Ontivero Ortega, Michell Valdés Sosa, Hichem Sahli

## Abstract

**Antecedent:** The diagnosis of schizophrenia could be enhanced with objective neurophysiological biomarkers, such as the event related potential features in conjunction with machine learning procedures. A previous work extracted features from event related responses to three oddball paradigms (auditory and visual P300, and mismatch negativity) for the discrimination of schizophrenic patients. They used several classifiers: Naïve Bayes, Support Vector Machine, Decision Tree, Adaboost and Random Forest. The best accuracy was obtained with Random Forest (84.7%).

**Objective:** The aim of this study was to examine the efficacy of Multiple Kernel Learning classifiers and Boruta feature selection method exploring different features for single-subject classification between schizophrenia patients and healthy controls.

**Methods:** A cohort of 54 schizophrenic subjects and 54 healthy control subjects were studied. Three sets of features related to the event related potentials signal were calculated: Peak related features, Peak to Peak related features and Signal related features. The Boruta feature selection algorithm was used to evaluate its impact on classification accuracy. A Multiple Kernel Learning algorithm was applied to address schizophrenia detection.

**Results:** We obtained a classification accuracy of 83% using Multiple Kernel Learning classifier with the whole dataset. This result in accuracy triangulates previous work and shows that the differences between schizophrenic patients and controls are robust even when different classifiers are used. Appling the Boruta feature selection algorithm a classification accuracy of 86% was yielded. The variables that contributed most to the classification were mainly related to the latency and amplitude of the auditory P300.

**Conclusion:** This study showed that Multiple Kernel Learning can be useful in distinguishing between schizophrenic patients and controls. Moreover, the combination with the Boruta algorithm provides an improvement in classification accuracy and computational cost.

## 1 Introduction

Schizophrenia is a severe and persistent debilitating psychiatric disorder with prevalence of 1% of the world population (McGrath et al., 2004). Although psychotic symptoms such as hallucinations and delusions are frequently present, impaired information processing is probably the most common symptom (Javitt et al., 1993). This deficit is reflected mainly in deficits in attention and working memory tasks when compared with healthy controls (Li et al., 2018). The diagnosis of schizophrenia is made by psychiatrists by ascertaining the presence of predefined symptoms (or their precursors) with personal interviews. However, in some cases this diagnosis is unclear, or patients are misdiagnosed with Schizophrenia (Coulter et al., 2019). Thus, finding biomarkers for the prediction of individuals with schizophrenia would be desirable in order to choose the optimal treatment (pharmacologic or non-pharmacologic). Analysis of EEG recording during information processing tasks could provide objective complimentary measures to support the subjective human-based decision process (Sabeti et al., 2009; Koukkou et al., 2018).

EEG is a non-invasive and low-cost technique used to measure electrical brain activity along multiple scalp locations. EEG signals have been widely adopted to study mental disorders, such as dementia, epileptic seizures, cognitive dysfunction, among others, as well as schizophrenia (Loo et al., 2016; Olbrich et al., 2016; Horvath et al., 2018). EEG reflects the spontaneous activity of myriad brain parcels, but also can include responses to afferent stimuli (Cong et al., 2015). Event related potentials (ERPs) are electrical responses that are time-locked to a specific stimulus or event, and can be used to assess brain dynamics during information processing in specific tasks (Woodman, 2010). When a subject is presented with a series of standard stimuli, interspersed with infrequent deviant stimuli, the Mismatch Negativity (MMN) (Lee et al., 2017) and the P300 (Li et al., 2018) components are generated. This task is known as the oddball paradigm and is used to study schizophrenia since consistent deficits in the P300 and MNN have been reported in this disease (Bramon et al., 2004; Javitt et al., 2017). Although MMN and P300 are usually produced by an infrequent unexpected event in a sequence of auditory stimuli, P300 can also be obtained with visual stimuli. The MMN is of shorter latency and does not require attention to the stimulus (Näätänen et al., 2004), whereas the P300 is of longer latency and requires attention to the stimulus (Huang et al., 2015).

Several studied have reported significant differences in the latency and amplitude of MMN and P300 between controls and patients, suggesting that these features are possible markers of the prodromal phase of schizophrenia (Atkinson et al., 2012; Loo et al., 2016) as well as a potential endophenotypes for schizophrenia (Earls et al., 2016). Analysis of a large dataset of auditory P300 ERP (649 controls and 587 patients) confirmed the reliability of this reduced amplitude, with a large effect size (Turetsky et al., 2015). However, these findings of statistically significance differences in a group analysis does not imply that EEG is useful for the prediction of individual schizophrenia cases (Lo et al., 2015), which requires applying a prediction paradigm using Machine Learning.

Accordingly, machine learning techniques are being applied to classify between schizophrenics (SZs) and healthy controls (HCs) using ERPs. The most common features used are based on amplitude and latency of different components (e. g. N100 and P300 (Neuhaus et al., 2013), P50 and N100 (Iyer et al., 2012; Neuhaus et al., 2014)), with several classifiers tested. Neuhaus et al. using visual and auditory oddball paradigms and a k-nearest neighbor (KNN) classifier obtained a classification accuracy of 72.4 % (Neuhaus et al., 2013). The same author with a bigger sample size and a Naive Bayes (NB) classifier achieved a 77.7% of accuracy (Iyer et al., 2012). Laton et al. evaluated the performance of several classifiers extracting features from auditory/visual P300 and MMN (Laton et al., 2014). The results using NB and Decision Tree (without and with AdaBoost) achieved accuracies of about 80%. Recently, Barros et al. published a critical review that summarizes machine learning-based classification studies to detect SZs based on EEG signals, conducted since 2016, (Barros et al., 2021). These authors reported that Support Vector Machines (SVM) were the commonly used algorithms, probably due to its computational efficiency. This kernel-based learning method also achieved the best performance in most studies. Nevertheless, none of the studies focused on ERPs, have used multiple kernels, employing instead only one specific kernel function.

The multiple kernel learning (MKL) method learns a weighted combination of different kernel functions and is able to benefit from information coming from multiple sources (Wani and Raza, 2018). It has been used to address the problem of biomarker evaluation for schizophrenia detection, but basically applied to Magnetic Resonance Images increasing performance accuracy (Ulaş et al., 2012; Castro et al., 2014; Liu et al., 2017). However, as far as we know, application of MKL to electrophysiological data has been not explored for schizophrenia, even though some authors are applying this technique to EEG signals for other purposes, mainly brain computer interfaces (Li et al., 2014; Zhang et al., 2017). Thus, MKL has not been applied in the objective diagnosis of Schizophrenia using EEG.

Here, using the same dataset provided by Laton et al. (Laton et al., 2014), we extended the set of predictor variables beyond the latency and amplitude of the ERP components, by including additional morphological features (based on time) together with some features extracted from the frequency domain. Due to the large number of features, the Boruta method was applied, which is a wrapper Random Forest (RF) based feature selection algorithm, to estimate the impact of a subset of important and relevant feature variables in the classification accuracy. The multiple kernel learning (MKL) was evaluated for the classification of SZs versus HCs.

## 2 Materials and methods

### 2.1 Dataset

The study was carried out on data from 54 patients and 54 controls, matched for age and gender. Patients were classified by a semi-structured interview (OPCRIT v4.0) and all participants gave written informed consent. Detailed demographic data can be found in **Table 1**. EEGs were recorded using a 64-channel and the international 10/10 system, with a sampling frequency of 256 Hz. Three paradigms auditory/visual P300 and MMN were used. **Table 2** shows a brief description of paradigms.

**TABLE 1.** Demographic data.

|  | Patients | Controls | P (t-test) |
| --- | --- | --- | --- |
| Number of participants | 54 | 54 |  |
| Male | 36 | 36 |  |
| Age (years): mean ± std | 40.5 ± 10.1 | 37.6 ± 14.1 | 0.22 |
| Age (years): range | [22.4, 60.5] | [15.1, 64.4] |  |
| Education (years): mean ± std | 12.6 ± 1.80 | 14.8 ± 2.11 | $4.84 \times 10^{-5}$ |
| Disease duration (years): mean ± std | 14.8 ± 9.04 | – | |
| Disease duration (years): range | [1, 40] | – |  |

**TABLE 2.**
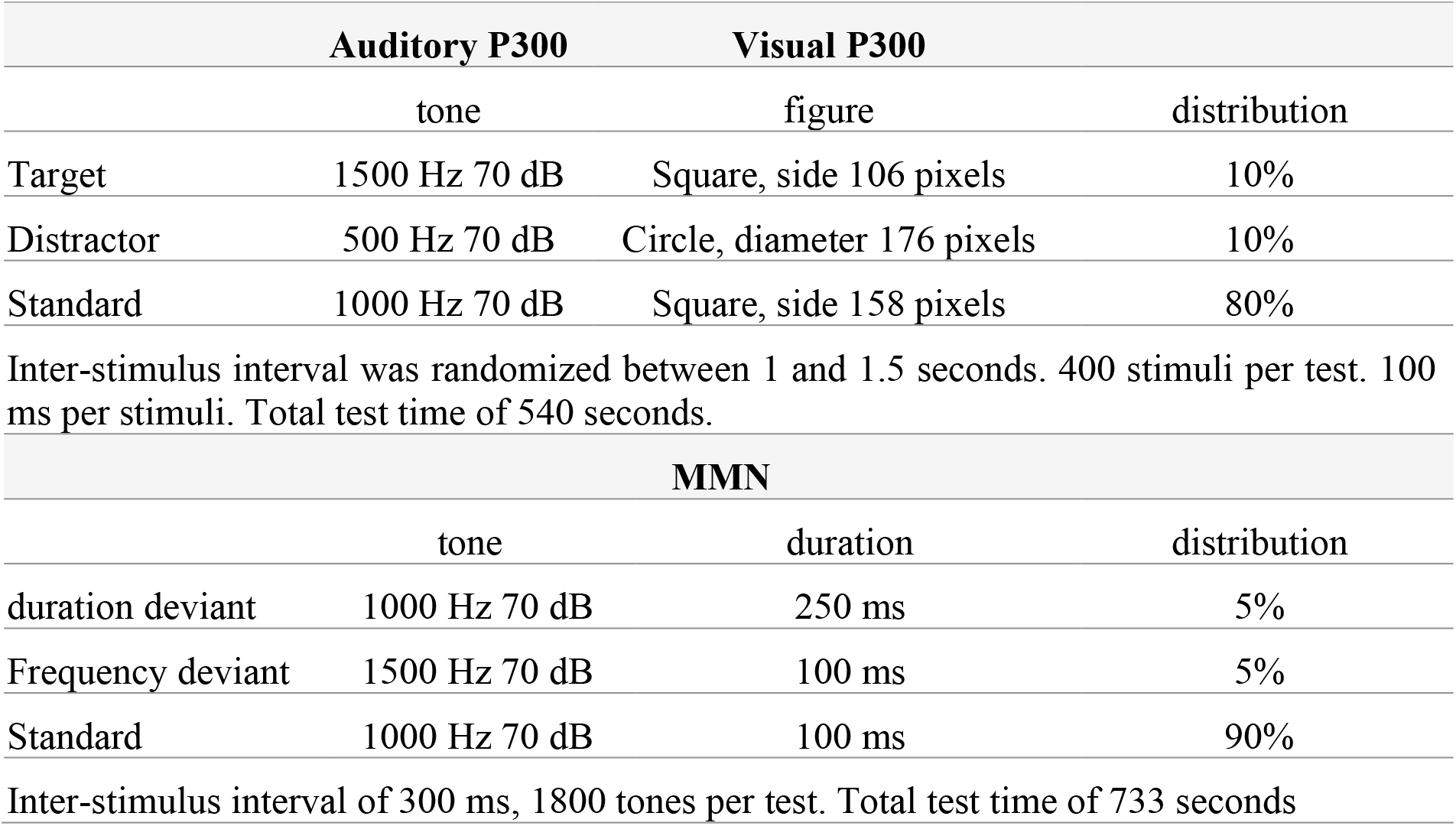
Paradigms and procedures

The signals were filtered using bandpass Butterworth filters with cuttoffs at 0.1 and 30 Hz. Epochs were extracted using time windows between -200 and 800 ms for the P300 paradigms, and between - 100 and 500 ms for the MMN. Subsequently, baseline correction, re-referencing to linked ears and artefact rejection were performed. Finally, epochs were averaged into stimulus specific responses for each individual and low-pass filter and baseline correction were re-applied. More details can be found in Laton et al (Laton et al., 2014).

### 2.2 Feature extraction

Feature extraction has been carried out on the waveform of ERPs emerged as the averaging of the electrical responses corresponding to the set of stimuli different from the standard stimulus (Target and Distractor for P300, Duration and Deviant for MNN)). Only Fz, Cz and Pz channels were considered (see **Figure 1**). Thus, the number of features extracted for classification purposes was 726 (282 features for each P300 paradigms and 162 for MMN paradigm). The feature values were standardized to ensure that all of them have equal weight during training of the classifiers. These standardized values were then normalized, rescaling them all to values between 0 and 1. In this binary classification problem, patients and controls were 1 and 0 respectively.

**FIGURE 1.**
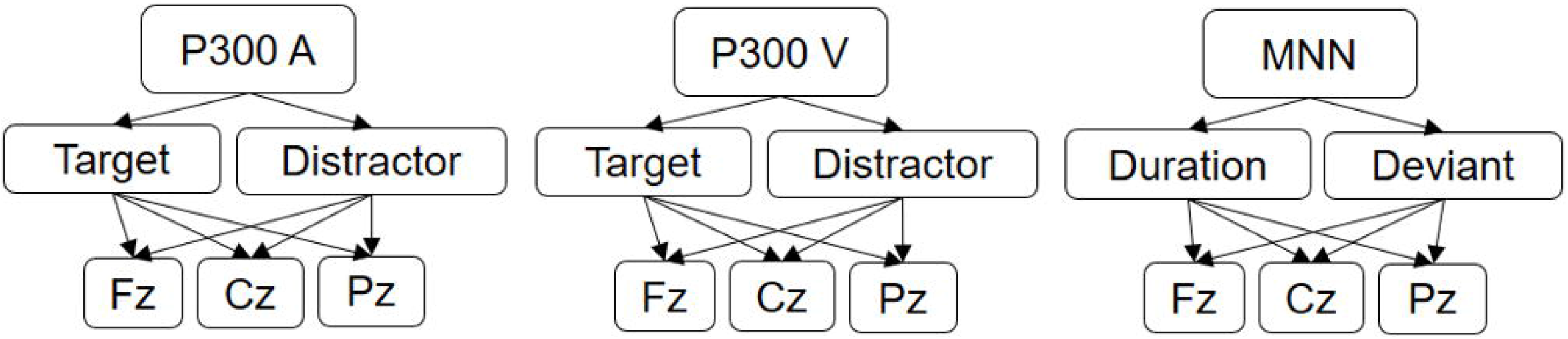
Averaged evoked potential signals used for feature extraction.

The set of features can be divided into three categories: Peak related features, Peak to Peak related features and Signal related features. Details about feature definitions are presented in **Annex 1**. Some of these features were previously used for other authors to calculate features related to the ERP signal (Kalatzis et al., 2004; Abootalebi et al., 2009). Four peaks for P300 paradigms (N100, P200, N200, and P300) and two peaks for MMN paradigm (N200, P300) were considered (see **Figure 2**).

**FIGURE 2.**
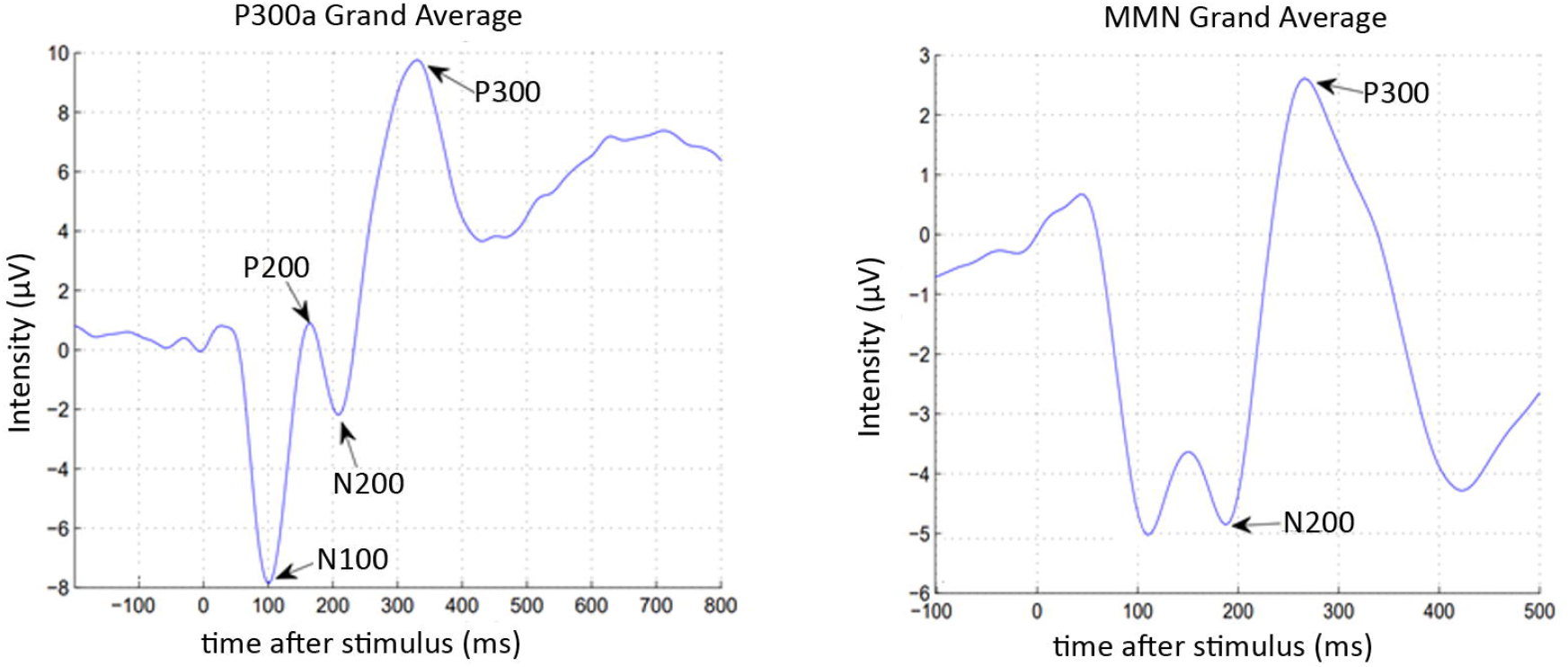
Principal components of P300 tasks (N100, P200, N200, P300) and MMN task (P200, P300).

#### 2.2.1 Peak related features

Peaks were estimated using the same algorithm described in Laton et al (Laton et al., 2014). The algorithm detects the largest absolute value in an interval established around the average latency of the peak in the respective grand average. This value is considered as *Amplitude* of the corresponding peak, their *Latency* is the time where the peak appears in the respective time interval. To ensure little overlap between the intervals, the detection interval was extended to contain the latency of peak most deviated. To search the latency of the peak, the minimum value of the corresponding detection interval was changed by the latency of the previous searched peak to avoid mistakes in the order of the ERPs components. The other features were: *Absolute Amplitude, Latency/Amplitude ratio, Absolute Latency/Amplitude ratio, Average Absolute Signal Slope* and *Slope sign alterations*.

#### 2.2.2 Peak to Peak related features

Three features were calculated considering the relationship between adjacent selected peaks: the absolute difference between the amplitude of the peak and the next peak in latency order; the difference in latencies of these two peaks; and the slope of the signal in this time window.

### 2.2.3 Signal related features

Features considering the area under the curve were calculated: the sum of the positive signal values (*Positive Area*); the sum of the negative signal values (*Negative Area*); the *Total Area*, and *Absolute Total Area*. Two more features related to the whole signal were calculated: the number of times that the amplitude value of the signal crosses the zero y-axis between two adjacent peaks (*Zero Crossing*); and the relation of the number of crosses per time interval (*Zero Cross Density*).

Additionally, frequency domain features were extracted using a Power Spectral Density (PSD) analysis: the frequency with the largest energy content in the signal (*Mode frequency*) spectrum; the frequency that separates the power spectrum into two equal energy areas (*Median frequency*); and an estimate of the central tendency of the derivate power distributions (*Mean frequency*).

### 2.3 Classifier used in the study

#### 2.3.1 MKL

The use of MKL has shown that it enhances the interpretability of decision functions and can improve classification performance compared with other classifiers (Kloft et al., 2009; Varma and Babu, 2009). Similar to simple SVM applications, this method is based on kernel definitions, however, instead of one single kernel, MKL combines several kernel functions (reflecting different kinds of information), and also automatically determines the importance of each kernel (Gönen and Alpaydin, 2011).

Given a set of data X and a feature mapping function Φ, a kernel matrix can be defined as the inner product of each pair of feature vectors:

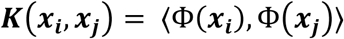

In the multiple kernel learning problem for binary classification, ***N*** data points **(*x***_***i***_, ***y***_***i***_**) (*y***_***i***_***ϵ* ± 1)** are given, where ***x***_***i***_ is translated via ***M*** mappings 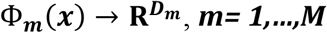, from the input into ***M*** feature spaces **Φ**_**1**_**(*x***_***i***_**), …**., **Φ**_***M***_**(*x***_***i***_) where ***D***_***m***_ denotes the dimensionality of the ***m***^***th***^ feature space.

Multiple Kernel Learning methods aim to construct an optimal kernel model where the kernel is a linear combination of fixed base kernels. Learning the kernel then consists of learning the weighting coefficients **β** for each base kernel, rather than optimizing the kernel parameters of a single kernel.

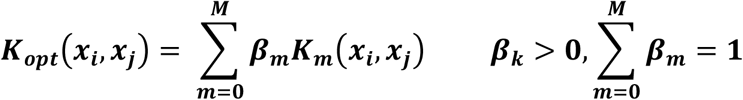

When MKL is plugged into SVM, the primal form of MKL is reformulated as the following optimization problem:

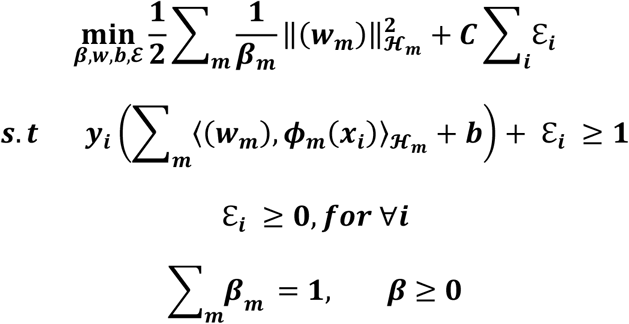

where ***C*** is a regularization parameter between training errors and an optimal separating hyperplane. For binary classification MKL problem, optimization is solved using semi-infinite programming (Sonnenburg et al., 2006). The three commonly used kernels are: linear kernel **(*K***_***L***_), polynomial kernel **(*K***_***P***_), and Gaussian kernel (***K***_***g***_):

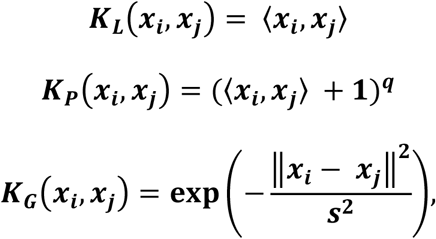

where parameter ***q*** is the polynomial degree and parameter ***s*** determines the width for Gaussian distribution.

MKL provides a general framework for learning from multiple and heterogeneous data sources (de Carvalho, 2019). This machine learning algorithm works by first constructing a kernel from each of the data sources and then combining these kernels based on a certain criterion for improved classification performance. With ***M*** kernels, a given input data can be mapped into ***M*** feature spaces. Another approach is when different basis kernels are applied to the same data features to identify which kernel is best for the problem at hand.

In this paper, the input data was mapped into different feature spaces trying to group variables with common aspects: type of paradigm, channels (Fz, Cz, Pz), or type of feature. For every feature space the 726 features were rearranged in three groups considering the common aspects (see **Figure 3**). Then, the MKL available in SHOGUN toolbox was applied (Sonnenburg et al., 2010) for every feature space. We used a non-sparse MKL with L2-norm that have more advantages over sparse integration method for thoroughly combining complementary information in heterogeneous data sources. L2-norm distributes the weights over all kernels while taking advantages of the effects of kernels in the objective optimization (Yu et al., 2010).

**FIGURE 3.**
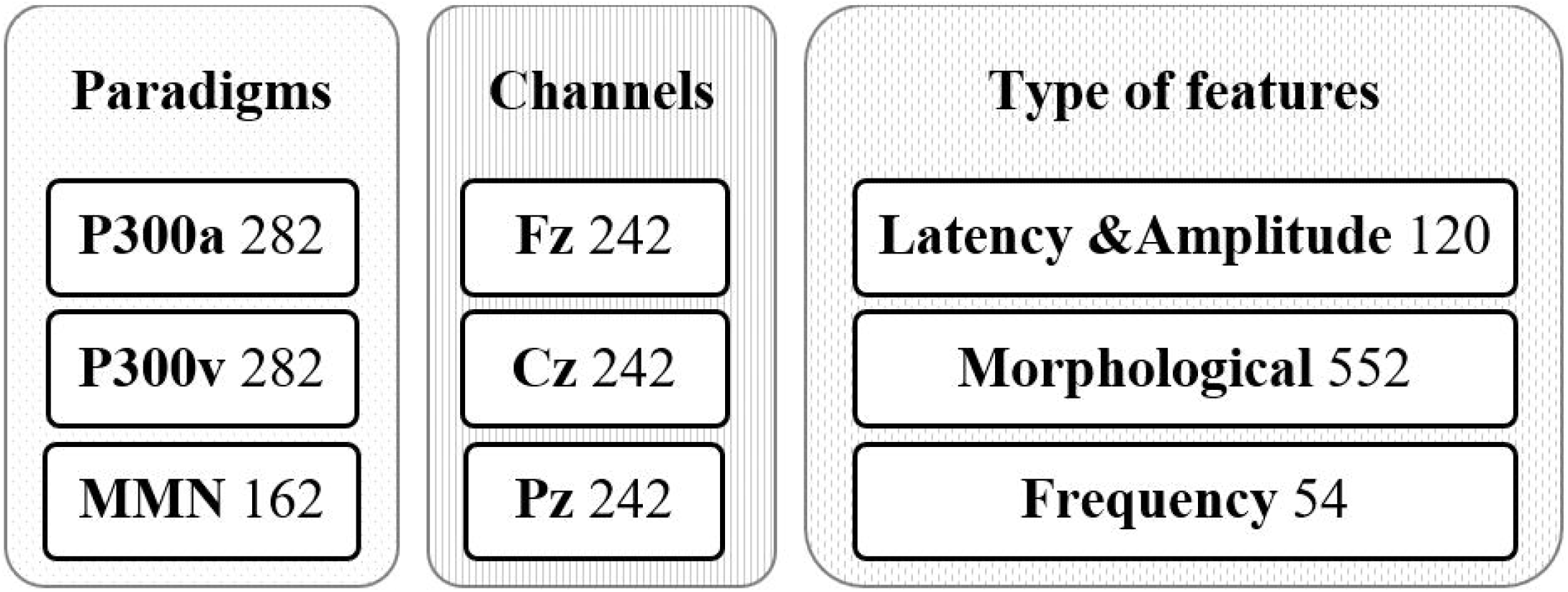
Grouping input data (726 features) in three possible kernel combinations according to the feature space approach.

### 2.4 Feature Selection

Feature selection yields a subset of features from the original set of features, which are the best representatives of the data. Therefore, it allows us to reduce the number of input variables. The goal of this process is to reduce the computational cost when developing a predictive model and, in some cases, to improve the performance of the model, not always guarantee (Benouini et al., 2020).

### 2.5 Boruta algorithm

Boruta is a feature selection algorithm that uses a wrapper method based on the RF classifier to measure the importance of variables. RF makes it relatively fast due to its simple heuristic feature selection procedure (Kursa, 2017). In the Boruta algorithm, the original feature set is extended by adding shadow variables (Kursa and Rudnicki, 2010). A shadow variable is created by shuffling values of the original feature. The importance values are calculated for all the attributes by running RF classifier resulting in a *Z score*. The maximum *Z score* is calculated among those shadow variables to assign a hit for each feature that scored better than this maximum. A two-sided test of equality is performed to obtain a statistically significant division between relevant and unimportant feature variables. If a variable systematically falls below the shadow ones, its contribution to the model is doubtful and is therefore eliminated. The shadow variables are removes and the process continues until all variables are accepted, rejected or a limit number of iterations is reached. This limit corresponds to the maximal number of RF runs.

The package “Boruta” in R was used (Kursa and Rudnicki, 2020). The implementation defaults to 100 as the maximum number of RF runs. To get a reduced number of attributes left undecided, this value was set to 500. Nevertheless, when this value isn’t enough, another function *TentativeRoughFix*, contained in the package, can be used to analyses those attributes which importance is very close to the decision criteria.

### 2.6 Nested cross validation

For explore the feature selection impact, nested cross validation (NCV) was applied. The NCV is characterized by having an inner loop responsible for model selection/hyperparameter tuning and an outer loop is for error estimation. The entire data was divided randomly into ***k*** subsets or folds with stratification, the same proportion of patients and controls as in the complete dataset. The ***k-1*** subsets are used for feature selection and the remaining subset for testing the model after feature selection. As in k-fold cross-validation method, this process was repeated ***k*** times (outer loop), each time leaving out one of the subsets reserved for testing and the rest for feature selection using Boruta algorithm (see **Figure 4**).

**FIGURE 4.**
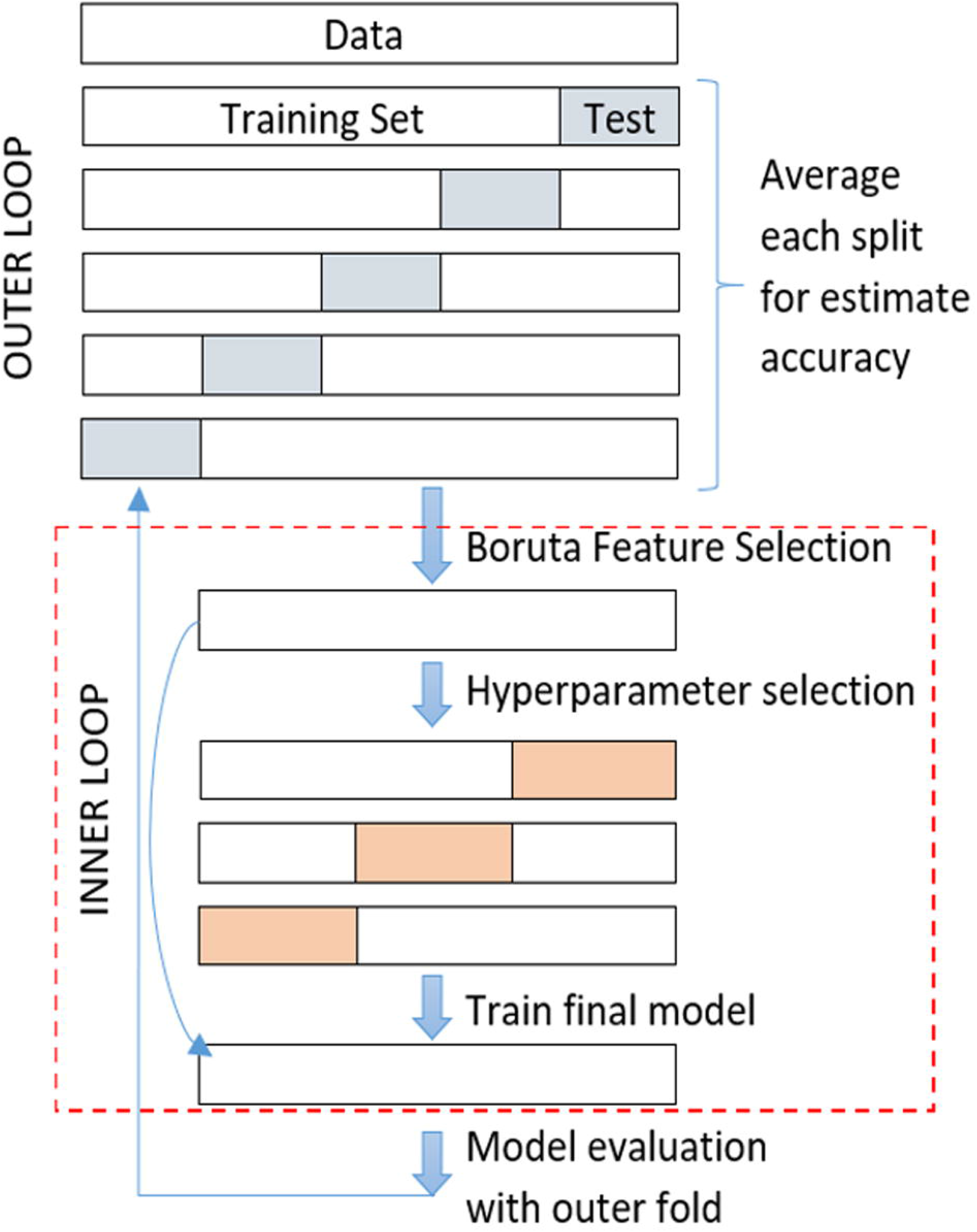
Feature selection steps applying nested cross validation.

Each subset obtained after feature selection, was used for model hyperparameter tuning in the inner loop. One of the approaches commonly used in practice for the selection of hyperparameters is to try several combinations of them and evaluate their out of sample performance. The tuned parameters in the MKL classifier were:

- Regularized parameter ***C***, a tradeoff between misclassification and simplicity of the model, the candidate’s values for grid was 0.5, 1, 1.5, 5, 10
- Type of kernel (linear, RBF, and polynomial)
- In case of RBF kernels the Sigma (***σ***) to determine the width for Gaussian distribution, exploring the following values 10, 5, 1, 0.25, 0.5, 0.75.

The parameter configuration selected to train the final model was the one that reached the highest average accuracy on the inner loop. The whole dataset used for tuning parameters was then trained and tested with its corresponding test set in the outer loop. The classifiers’ performance was obtained by averaging the accuracy of the ***k*** trained models.

## 3 Results

### 3.1 Feature Selection

The Boruta algorithms yielded an average of 32 attributes selected per ***k*** iteration with values in a range of 26 to 42 (see **Figure 5A**). The median computation times was around 2.6 minutes (std 0.04), with 0.005 min per RF runs. A total of 76 attributes were selected at least once. **Figure 5B** shows how many times these attributes were selected in the process. The distribution of variable per paradigm is also showed. The 80% of the 76 attributes selected were related to amplitude, latency, or the correlation between them. Attributes related to frequency domain was barely selected.

**FIGURE 5.**
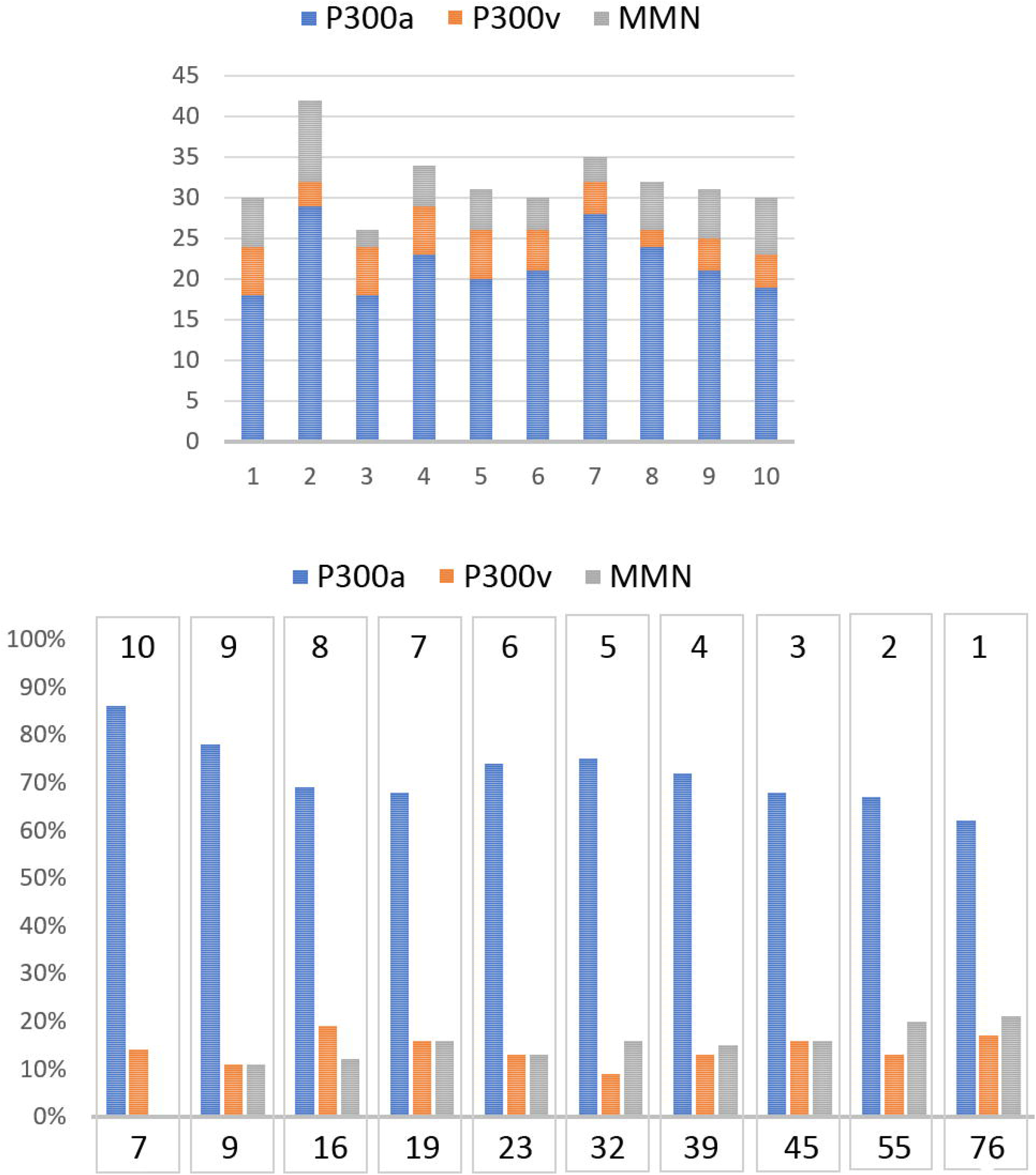
Distribution of feature selection in 10-fold-cross-validation. **(A)** Distribution per paradigm in the 10 subsets of features selected. **(B)** Frequency of selection of all the attributes that were selected in the ten Boruta applications. The bottom number means how many features were selected the number of times represented in the top number.

Only seven features were identified as important every time Boruta algorithm was used. **Table 3** describes these features according to the paradigm, type of stimulus, channel, and type of feature.

**TABLE 3.** Features always selected by Boruta

| PARADIGM | STIMULUS | CHANNEL | PEAK | FEATURE |
| --- | --- | --- | --- | --- |
| P300v | Target | Pz | P2 | latency |
| P300a | Distractor | Cz | N1 | absRatio |
| P300a | Distractor | Fz | P2 | absRatio |
| P300a | Distractor | Fz | P2 | absAmplitude |
| P300a | Target | Cz | N1 | absRatio |
| P300a | Target | Cz | N2 | latency |
| P300a | Target | Cz | P2 | latency |

### 3.2 Classifier performance

To compare the performance of the MKL algorithms three metrics derived from the confusion matrix were used. As the classes were balanced, accuracy (*Acc*) is a good measure for assessing the classification models. Accuracy is the proportion of the total number of predictions that were correct. The other two measures were sensibility (*Sen*) that evaluates true positive rates, and specificity (*Spe*) to evaluate the false positives rates. In **Table 4**, the performance of MKL algorithm when feature selection was applied or not is shown.

**Table 4.** Performance (%) of MKL algorithm with and without Boruta feature selection

| MKL Kernels | Without FS |  |  | With FS |  |  |
| --- | --- | --- | --- | --- | --- | --- |
|  | Acc | Sen | Spe | Acc | Sen | Spe |
| Paradigm | 83 | 80 | 88 | 86 | 86 | 87 |
| Channels | 80 | 74 | 87 | 84 | 85 | 86 |
| Type of Features | 82 | 78 | 85 | 86 | 86 | 86 |

### 3.3 Discussion

Here we explored the use of MKL classification algorithm for distinguishing SZs from HCs based on ERP data. Using all features, the best classification accuracy (83%) was achieved when kernels were built by grouping features according to paradigms. Moreover, when MKL was combined with Boruta method, a classification accuracy of 86% was obtained. With this feature selection algorithm, the large number of predictor variables was reduced significantly (96%) with a lower computation time. Therefore, training time of MKL was also reduced, its main shortcoming is known to be its high computational cost, especially when many features are used (de Carvalho, 2019).

Review of the Boruta algorithm results pointed out that variables with major importance were mainly related with auditory P300 ERP paradigm. This correspond with the general finding that the P300 measures obtaining from auditory stimuli are more effective in differentiating SZs from HCs than those obtaining from visual stimuli (Park et al., 2005). An interesting point to be noted is that feature selected by Boruta were mainly related with amplitude, latency, and correlation between them. These features correspond with Peak related features. To a lesser extend Peak to Peak related features was included in the selection. However, only three features of Signal related features were rarely included, thus features in frequency domain didn’t contribute to classification.

Overall, these findings are in accordance with findings reported by other authors, and thus triangulates the previous results and shows that the differences between SZs and HCs are robust even when different classifiers are used. Numerous authors have been concluded that odd-ball tasks are potential biomarker for diagnosis in schizophrenia. Some of them have verified that the use of latency and amplitude produces similar results in the discrimination of SZs from HCs. Santos-Mayo et al. used time and frequency ERP features, they explored several electrodes grouping, classifiers, feature selection algorithms and filtering schemes (Santos-Mayo et al., 2017). They achieved accuracies above 90% but their dataset was unbalanced and small, which could limit the generalization of their findings. Shim et al. proposed to extend P300 amplitude and latency sensor-level feature with cortical current density values as source-level features, due to the low spatial resolution originating from volume conduction (Shim et al., 2016). Using Fisher’s scores, feature set ranged for 1 to 20 were selected for classification. They reported classification accuracies of 81% for sensor-level features, 85% for source-level features and 88% combined them, using SVM classifier. Laton et al. combined latency and amplitude features of responses to three different odd-ball tasks to apply several classification algorithms (Laton et al., 2014). They achieved a classification accuracy averaged 77% (3.5 std) and their best result, closed to 85%, corresponded to RF classifier. These authors also found a similar pattern in terms of the most relevant features, since in a ranking of the 20 main variables, 14 were extracted from the P300 auditory oddball paradigm. They stated auditory P300 as the most valuable of the three ERP paradigms to the final prediction success.

Compared with these previous studies, our accuracies values are in a range considered as a good accuracy, very close to the results previously reached. This result adds robustness to the previous findings remarking the possibility of accurately distinguish SZs from HCs using neurophysiological measurements. The present finding confirms that Boruta algorithm is a computationally efficient and robust algorithm that improves classification accuracy in many scenarios (Speiser et al., 2019).

Although the approach used here meet our goals, the information of the spatial voltage distributions over the scalp surface was wasted. It is known that the topography across the scalp was significantly different between schizophrenia and normal control groups (Morstyn et al., 1983; Frantseva et al., 2014). Some authors had investigated the topographic abnormalities of schizophrenia mainly group-based researches (Basile et al., 2004). However, individual patient-level analysis using topographic features has been less explored for schizophrenia. This would be a fruitful area for further work in other to reliably classify SZs from HCs.

This study suffers of small sample size as usual in psychiatric cohorts. In these cases, instead of a-priori train/validate/test partitions, strategies of cross-validation allow to estimate the selected model performance and avoid the risk of data leakage. Nevertheless, larger sets yield a more stable, reliable estimate of future performance and guarantee better generalization (Cearns et al., 2019).

## Data Availability

All data produced in the present work are contained in the manuscript

## 6 Acknowledgments

The authors would like to thank teams of Cuban Neuroscience Center and the Department of Electronics and Informatics (ETRO) of Vrije Universiteit Brussel (VUB) for supporting this research project.

## 7 ANNEX

**Annex 1:**
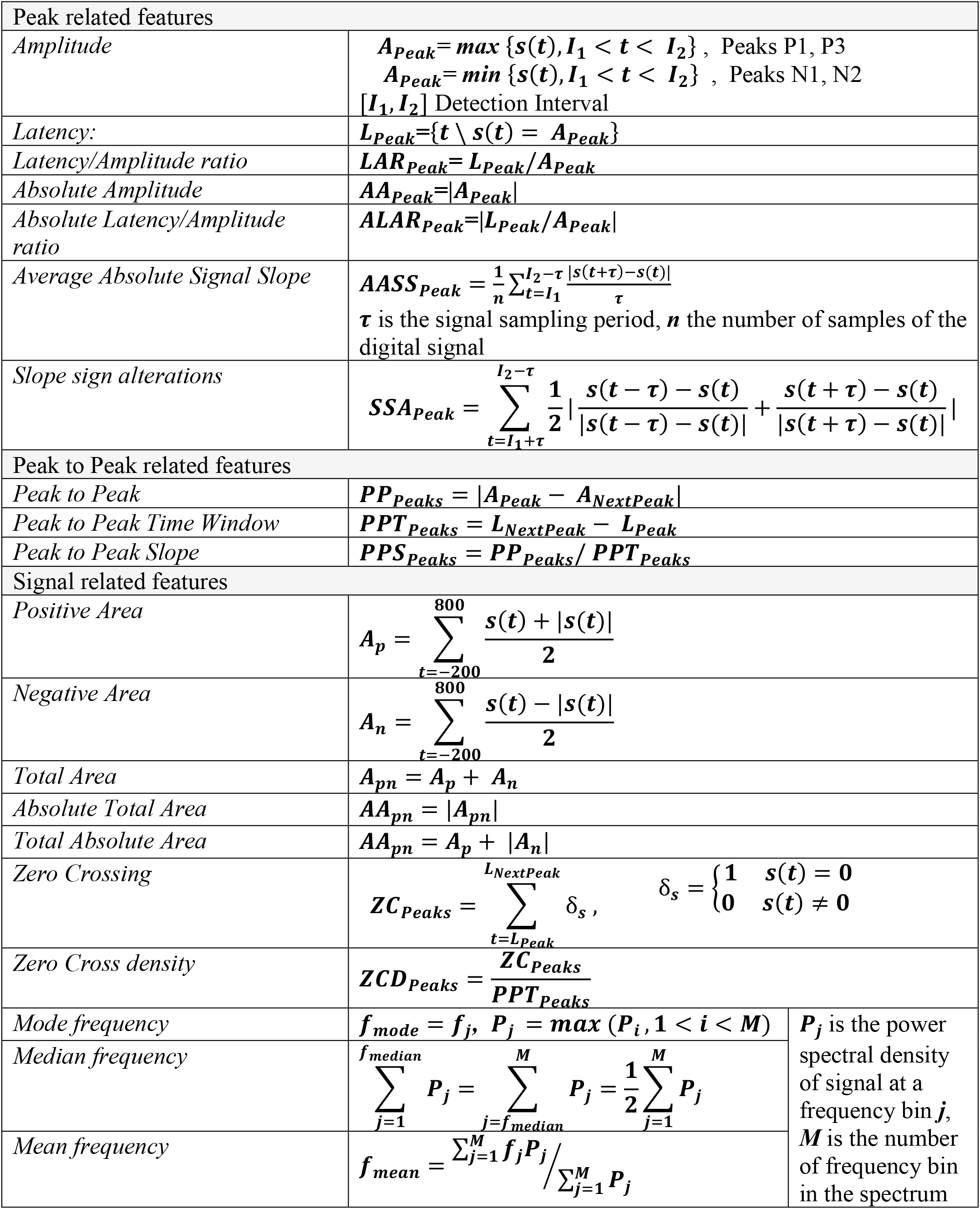
Feature definitions

